# Emergency department utilization and hospitalizations for ambulatory care sensitive conditions among unattached people actively seeking a primary care provider during the COVID-19 pandemic: a retrospective cohort study

**DOI:** 10.1101/2022.01.31.22270015

**Authors:** Emily Gard Marshall, David Stock, Richard Buote, Melissa K. Andrew, Mylaine Breton, Benoit Cossette, Michael E. Green, Jennifer E. Isenor, Maria Mathews, Anders Lenskjold, Adrian MacKenzie, Ruth Martin-Misener, Beth McDougall, Melanie Mooney, Lauren R. Moritz

## Abstract

**Background:** Primary care (PC) attachment improves healthcare access and prevention and management of chronic conditions. Yet, growing proportions of Canadians are unattached, signing-up on provincial waitlists. Understanding variations in healthcare utilization during COVID-19, and among potentially vulnerable unattached patients, is needed. This study compares emergency department (ED) utilization and hospitalization among those on and off a provincial PC waitlist, during the first two waves of COVID-19.

**Methods:** Waitlist and administrative health data were linked to describe persons ever/never on the waitlist between January 1, 2017, and December 24, 2020. ED utilization and ambulatory care sensitive conditions (ACSC) hospitalization rates by current waitlist status were quantified from physician claims and hospitalization data. Relative differences during COVID-19 first and second waves were compared with the previous year.

**Results:** During the study period, 100,867 Nova Scotians (10.1%) were on the waitlist. Those on the waitlist had higher ED utilization and ACSC hospitalizations. ED utilization was higher overall for individuals ≥65 years and females; lowest during first two COVID-19 waves; and differed more by waitlist status for those <65 years. ED contacts and ACSC hospitalizations decreased during COVID-19 relative to the previous year, and for ED utilization this difference was more pronounced for those on the waitlist.

**Interpretation:** Nova Scotians seeking PC attachment utilize hospital-based services more frequently than those not on the waitlist. Both groups had lower utilization during the COVID-19 pandemic than the year before. The degree to which forgone services produces downstream health burden remains to be seen.

## Introduction

In Canada, having a regular primary care provider is essential to efficiently accessing many publicly-funded health services.(1,2) Having a regular provider is associated with more effective preventative care, disease management, and coordination of care across systems leading to better health outcomes.(2– 4) Unfortunately, in 2020, roughly 10% of Canadians reported being “unattached” (i.e., not having a regular primary care provider or practice), which was among the worst when compared to peer countries.(5) To address this ongoing issue of access to care for unattached patients, several provinces created centralized waitlists.(2) In Nova Scotia (NS), this waitlist, launched in 2016, is known as the Nova Scotia Health *Need a Family Practice Registry*, holding data on registrant characteristics and health card identifiers facilitating linkage with administrative healthcare utilization data.(6) Throughout the COVID-19 pandemic, the publicly-reported number of registrants on the centralized waitlists has continued to grow.(7)

Having a significant proportion of unattached patients in the population has implications across the healthcare system. With limited alternatives, unattached patients seek care from walk-in clinics and visit emergency departments (EDs) for health concerns normally addressed within a primary care setting.(3,8) Inadequate access to primary care can lead to preventable hospitalizations, particularly for certain previously identified conditions, known as Ambulatory Care Sensitive Conditions (ACSC).(4) Low acuity ED visits and hospitalizations for ACSC are an inefficient use of health system resources and result in poorer patient and system outcomes.(4,9,10)

During a pandemic, ED utilization is expected to differ from usual patterns due to changes in health system policy, public safety concerns, and emerging health issues related to the pandemic. Patients may avoid visiting the ED due to fear of infectious disease transmission (11), or alternatively, may seek primary care in the ED or experience ACSC hospitalizations due to restricted access to community-based primary care providers.(12) As such, it is important to understand whether ED utilization and ACSC hospitalization rates differ for people with and without primary care attachment, particularly during the COVID-19 pandemic. NS recently reached a population of 1 million people, and the province is home to an older than average population who report more difficulty accessing after-hours care other than EDs(4,13). The objectives of this study are to describe ED utilization and ACSC hospitalizations among Nova Scotians who were either on or off a centralized primary care provider waitlist (hereafter referred to as “on-” or “off-Registry,”) and assess how utilization changes during the first and second waves of COVID-19.

## Methods

This study uses a descriptive cohort design to estimate population-based rates of ED utilization and ACSC hospitalizations among Nova Scotians identified as either formally seeking or not seeking a primary care provider based on quarterly “on-” or “off-Registry” status. The target underlying cohort comprises all publicly insured Nova Scotians ≥5 years as of April 1, 2016. The study period spans January 1, 2017, through December 24, 2020. Participants are considered if they have one or more days of enrollment within a calendar quarter.

This work is part of the Problems Coordinating and Accessing Primary Care for Attached and Unattached Patients in a Pandemic (PUPPY) study, funded by the Canadian Institutes of Health Research. The complete protocol for this study has been published previously.(14)

## Data Sources

This study used a novel linkage between centralized primary care provider waitlist data and administrative health data at Health Data Nova Scotia (HDNS). Linked administrative data holdings comprise:

- HDNS Insured Patient Registry, which identifies all publicly insured Nova Scotians eligible to receive primary care and contains demographic data such as age and sex;
- physician billings;
- Canadian Institute for Health Information (CIHI) Discharge Abstract Database (DAD).

Physician billings and the Discharge Abstract Database were used to estimate the Charlson Comorbidity Index.(15) Additional demographic measures, including after-tax household income, rurality of residence and the Canadian Index of Multiple Deprivation (CIMD)(16), are obtained by postal code-linked census (2016 Canadian Census) data.

**Key Measures**

To quantify ED utilization, ED contacts were captured from physician billing records that coded the ED as the hospital unit where the service was provided. Where identified, multiple records per date per patient were enumerated in analyses. From the initial admission date and throughout the duration of hospitalization, ACSCs were identified using ICD-10 diagnostic codes for seven condition clusters: epilepsy, chronic obstructive pulmonary disease, asthma, diabetes, heart failure and pulmonary edema, hypertension, and angina. These comprise the core set of conditions identified by CIHI (17) for which hospitalizations were deemed avoidable given provision of timely and effective outpatient care, either by avoiding condition onset, controlling the illness episode, or chronic disease management.(18)

The Charlson Comorbidity Index is derived by the weighted summation of specific comorbid conditions and was originally used to predict one-year inpatient mortality risk.(15) It has been adapted and weighted for use with Canadian administrative health data,(19) for outpatient populations,(20) and validated for comorbidity adjustment.(21) Rurality and after-tax household income were estimated using postal code-linked 2016 Canadian Census data contained in the Canada Post Postal Code Conversion File Plus.(22) Rurality is inferred by a community size of <10,000 people. The CIMD measures deprivation and marginalization across four dimensions: residential instability, economic dependency, ethnocultural composition, and situational vulnerability. Factor analysis-derived dimension-specific indices were created from selected Canadian Community Health Survey items and provide national and regional scores (i.e., Atlantic region used for this study).(23) Scores are provided at the level of the census dissemination area using 2016 Canadian Census Data. CIMD scores were summed and divided by four to produce an overall summary score.

### Analysis

Measures of central tendency for age and Charlson comorbidity index, and proportions for all demographic measures, were calculated for the entire NS primary care-eligible population, and by ever and never “on-Registry” status. We calculated “On-” and “off-Registry” ED utilization and ACSC hospitalization rates, the quarterly denominators of which were drawn from the NS primary care-eligible population, with replacement, for each interval. Chi-squared tests were used to assess differences in proportions across those ever “on” - and “off-Registry”. Unadjusted negative binomial regression was used to assess relative differences in ED and ACSC hospitalization rates across those “on-” and “off-Registry” by quarter. Rate ratios were estimated using generalized estimating equation approximations to multivariable negative binomial regression (to accommodate participants contributing to both intervals comprising comparison) comparing Q2 2020 (i.e., corresponding to COVID-19 1^st^ wave in NS) to Q2 2019 for ED utilization and ACSC hospitalizations and Q4 2020 (COVID-19 2^nd^ wave in NS) to Q4 2019 for ED utilization only (due to CIHI-DAD data access only to July 2020).

### Results

Table 1 describes the characteristics for the overall study population and by ever “on-Registry” status. There were 990,655 Nova Scotians ≥5 years of age as of April 1, 2016 identified in the HDNS Insured Patient Registry. Of these, 100,867 Nova Scotians were identifiable as ever “on-Registry” and were enrolled at least one day between January 1, 2017, and December 24, 2020. Proportions of individuals 50 years or younger and 80 years or older were smaller for people ever “on Registry,” and the proportion of females ever “on-Registry” was greater than the proportion of males. A nonzero Charlson comorbidity index, indicating at least one eligible comorbid condition, was more frequent among people “on Registry”. Rural Nova Scotians and people among the lower four aggregated household income categories were more frequently “on-Registry”. In contrast, people with the lowest level of deprivation were “on Registry” less frequently. (Above differences in proportions statistically significant at an alpha level of <0.0001)

**Table 1.** Description of NS “Primary Care User-eligible” cohort: overall; ever on/never on the Nova Scotia Need a Family Practice Registry centralized primary care provider waitlist

|  | NS Primary Care Population |  | Ever “on-Registry” |  | Never “on-Registry” |  |
| --- | --- | --- | --- | --- | --- | --- |
|  | n | mean (SD)/<br>% | n | mean (SD)/<br>% | n | mean (SD)/<br>% |
| **Overall** | 990655 |  | 100867 |  | 889788 |  |
| **Age** |  |  |  |  |  |  |
| Age (mean; SD) | 990655 | 45.5 (22.1) | 100867 | 46.7 (20.9) | 889788 | 45.3 (22.2) |
| 5-18 yrs | 136542 | 13.78 | 11899 | 11.80 | 124643 | 14.01 |
| 19-49 yrs | 411216 | 41.51 | 39371 | 39.03 | 371845 | 41.79 |
| 50-59 yrs | 165820 | 16.74 | 19138 | 18.97 | 146682 | 16.49 |
| 60-64 yrs | 73735 | 7.44 | 9358 | 9.28 | 64377 | 7.24 |
| 65-69 yrs | 67155 | 6.78 | 8389 | 8.32 | 58766 | 6.60 |
| 70-74 yrs | 47562 | 4.80 | 5654 | 5.61 | 41908 | 4.71 |
| 75-79 yrs | 34111 | 3.44 | 3701 | 3.67 | 30410 | 3.42 |
| ≥80 yrs | 54514 | 5.50 | 3357 | 3.33 | 51157 | 5.75 |
| **Sex** |  |  |  |  |  |  |
| Female | 503699 | 50.85 | 54726 | 54.26 | 448973 | 50.46 |
| Male | 486956 | 49.15 | 46141 | 45.74 | 440815 | 49.54 |
| **Charlson index** |  |  |  |  |  |  |
| 0 | 639744 | 64.58 | 57886 | 57.39 | 581858 | 65.39 |
| 1 | 206328 | 20.83 | 26142 | 25.92 | 180186 | 20.25 |
| 2 | 71375 | 7.20 | 8560 | 8.49 | 62815 | 7.06 |
| 3 | 30296 | 3.06 | 3676 | 3.64 | 26620 | 2.99 |
| ≥4 | 42912 | 4.33 | 4603 | 4.56 | 38309 | 4.31 |
| **Rurality** |  |  |  |  |  |  |
| Non-rural | 632745 | 63.87 | 58256 | 57.76 | 574489 | 64.56 |
| Rural | 331228 | 33.44 | 41794 | 41.43 | 289434 | 32.53 |
| Missing | 26682 | 2.69 | 817 | 0.81 | 25865 | 2.91 |
| **Household Income** |  |  |  |  |  |  |
| Q1 (lowest) | 191293 | 19.31 | 20712 | 20.53 | 170581 | 19.17 |
| Q2 | 193994 | 19.58 | 20990 | 20.81 | 173004 | 19.44 |
| Q3 | 186475 | 18.82 | 19833 | 19.66 | 166642 | 18.73 |
| Q4 | 195222 | 19.71 | 20452 | 20.28 | 174770 | 19.64 |
| Q5 | 196989 | 19.88 | 18063 | 17.91 | 178926 | 20.11 |
| missing | 26682 | 2.69 | 817 | 0.81 | 25865 | 2.91 |
| **Canadian Index Multiple Deprivation: Overall score** |  |  |  |  |  |  |
| 1 - 2 | 109464 | 11.05 | 8877 | 8.80 | 100587 | 11.30 |
| >2 - 3 | 380107 | 38.37 | 38929 | 38.59 | 341178 | 38.34 |
| >3 - 4 | 415015 | 41.89 | 45530 | 45.14 | 369485 | 41.53 |
| >4 - 5 | 58419 | 5.90 | 6686 | 6.63 | 51733 | 5.81 |
| missing | 27650 | 2.79 | 845 | 0.84 | 26805 | 3.01 |

Figure 1 displays the identified NS primary care eligible cohort “on-Registry” over the study period, enumerated monthly. Enrollment surpassed 10,000 during the first quarter (Q1-2017) then increased through Q4-2018, peaking at just over 43,000 in November. Registrations declined to just under 35,000 in Q2-2020, in line with the first wave of active COVID-19 cases.

**Figure 1.**
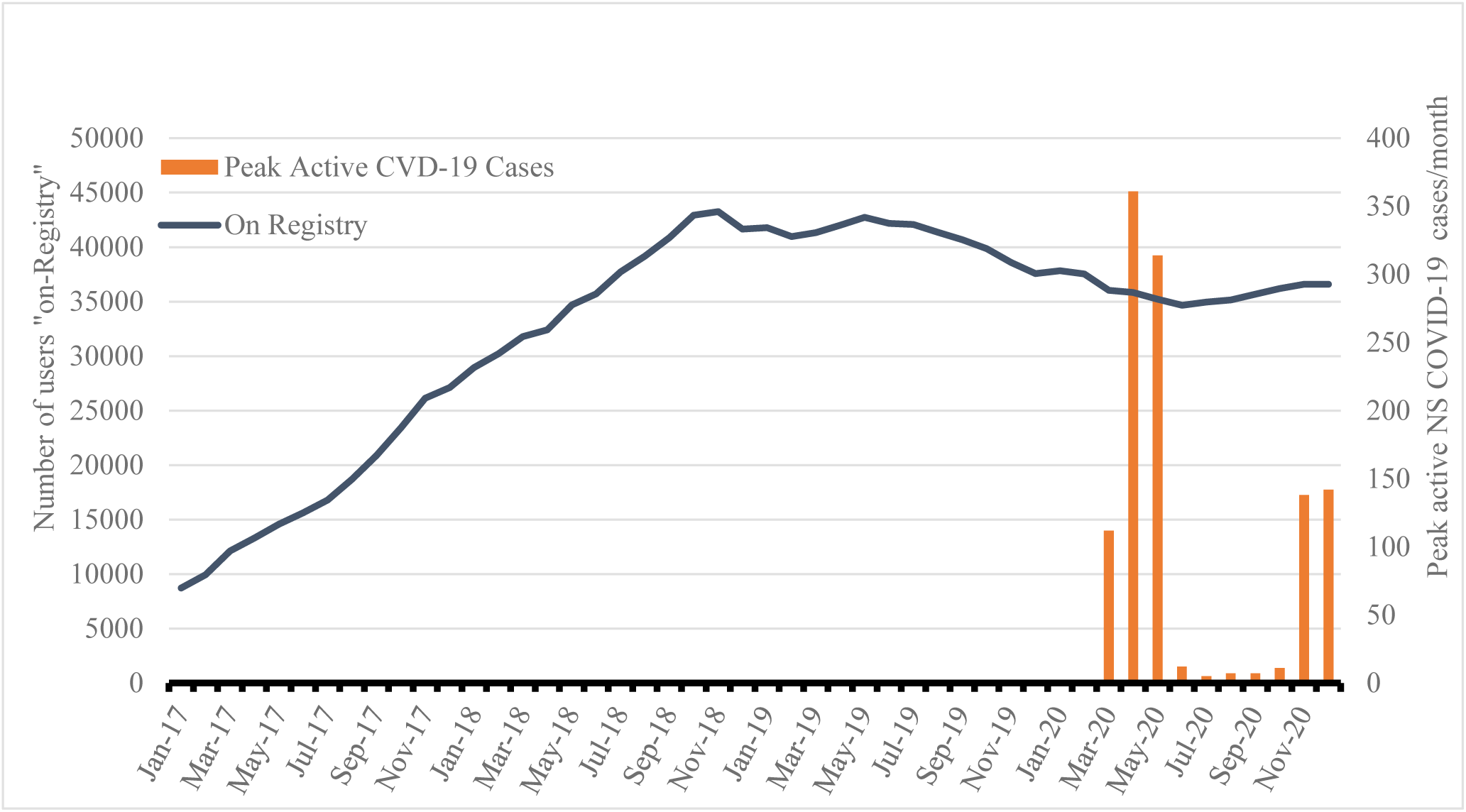
Number of Users “on-Registry” (CPW) by Month (Jan. 2017 – Dec. 2020)

Figure 2A shows overall rates of ED contacts. Aggregated over the entire study period, there were 155.9 and 105.3 ED contacts per 1,000 population among people “on-” and “off-Registry”, respectively. Individuals both “on-” and “off-Registry” had lowest rates during Q2- and Q4-2020, corresponding with NS’s COVID-19 first and second waves (CVD-19 Wave 1 and Wave 2, respectively) and utilization was consistently lower for those “on-Registry”. People ≥65 years had higher ED utilization rates (Figure 2B), though the difference between individuals “on-” and “off-Registry” was more pronounced among those <65 years (Figure 2C). While ED utilization was moderately higher for females (Figure 2D), both males (Figure 2E) and females “on-Registry” had higher utilization and achieved lowest utilization during COVID-19 first and second waves.

**Figure 2.**
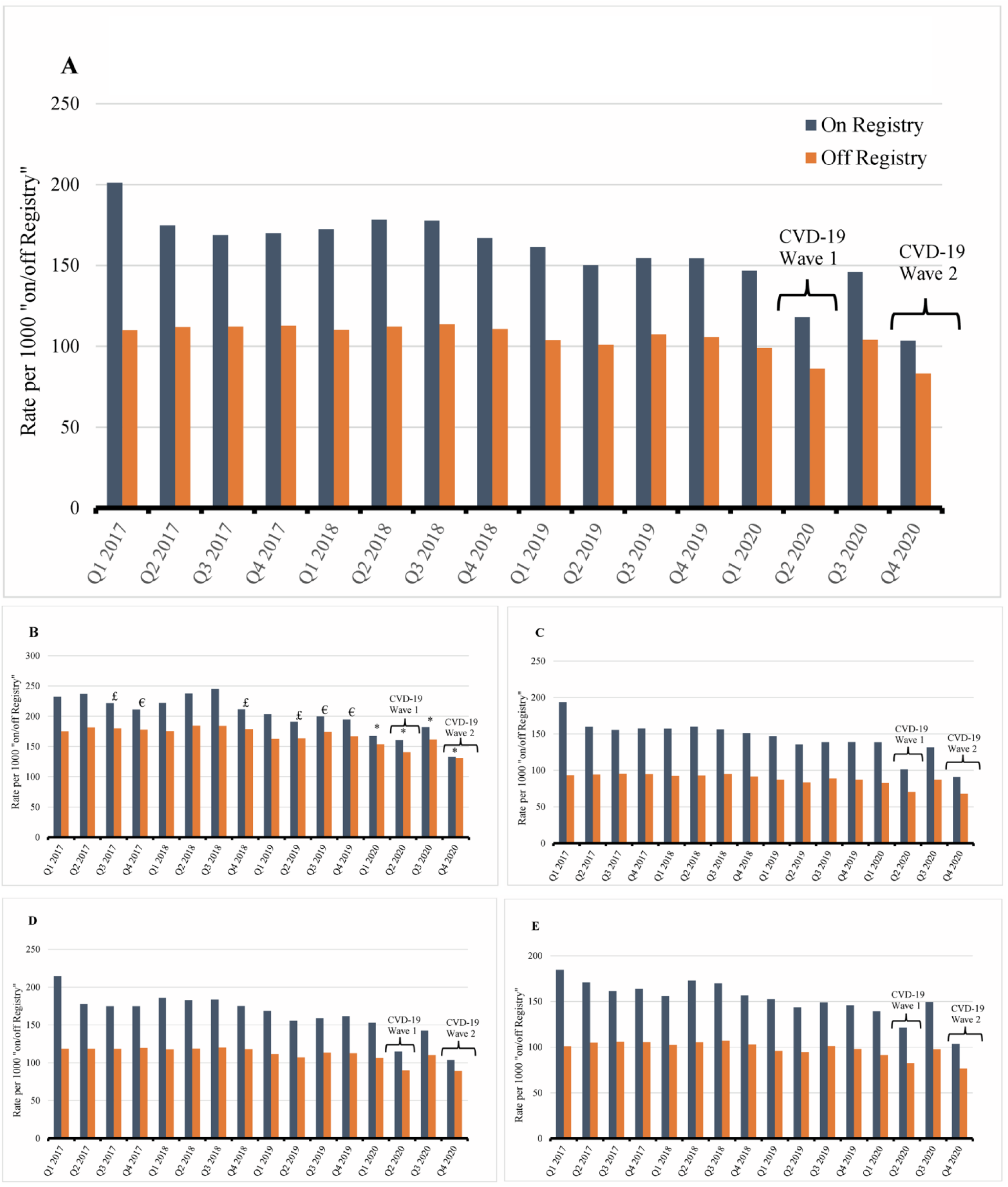
ED utilization (number of encounters), overall (A); ≥65 years (B); <65 years **(C)**; females **(D)**; males **(E)** Relative differences in rates statistically significant at p ≤0.01 unless specified; p ≤0.01 (£); p≤0.05(€); p>0.05 (*)

Overall, ACSC hospitalizations rates were higher for those “on-Registry” for most quarters (statistically significantly for six; Figure 3A). Similar to ED utilization, the lowest overall ACSC hospitalization rate (8.7 per 10,000 population) was observed during the COVID-19 first wave (Q2-2020) for those “off-Registry.” The highest ACSC hospitalization rates for people “on-Registry” occurred a year earlier in Q2-2019 (20.6 per 10,000 population).

**Figure 3.**
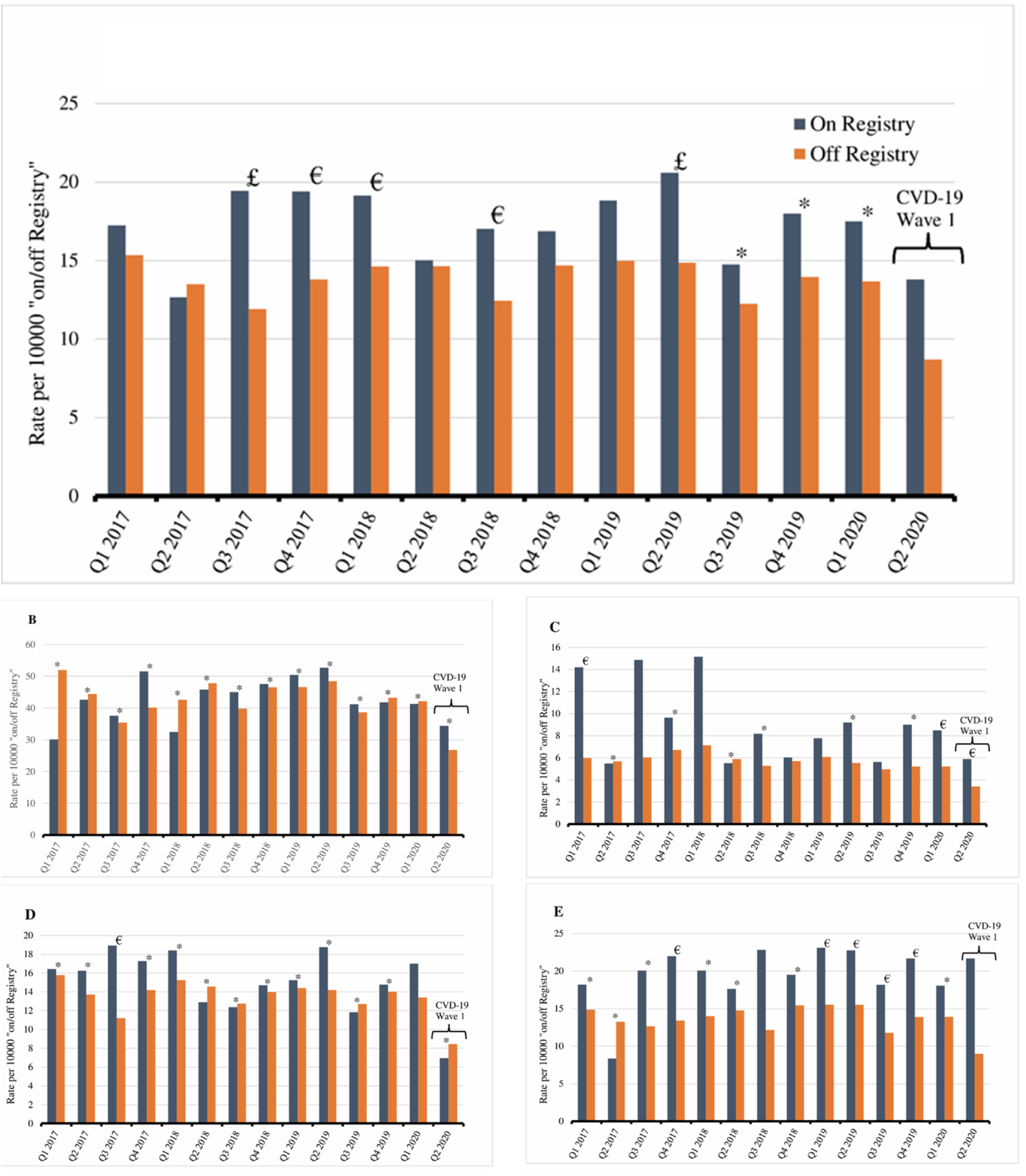
ACSC hospitalizations, overall **(A)**; ≥65 years **(B)**; <65 years **(C)**; females **(D)**; males **(E)** Relative differences in rates statistically significant at p ≤0.01 unless specified; p ≤0.01 (£); p≤0.05(€); p>0.05 (*)

Figure 3B shows quarterly ACSC hospitalization rates for those ≥65 years. Rates were relatively homogenous across registration status. The largest departure since Q1-2018 occurred during the COVID-19 first wave (Q2-2020), where it was higher for those “on-Registry”, though the difference was not statistically significant. Figure 3C shows ACSC hospitalizations for those <65 years. Rates for those “on-Registry” were higher for most quarters (statistically significantly for five), including COVID-19 first wave, though the outcome among younger Nova Scotians was relatively sparse.

Relative to males “off-Registry,” those “on-Registry” had routinely higher ACSC hospitalization rates from Q3-2017 onward (statistically significantly for seven; Figure 3E), including during COVID-19 wave one. Conversely, for females (Figure 3D), differences across registration status were attenuated.

Compared to the same quarter during the previous year (Table 2), ED utilization during the first wave of active COVID-19 cases in NS was moderately lower for both those “on-” and “off-Registry” (multivariable-adjusted “on-Registry” IRR: 0.86, 95% CI: 0.81-0.92; “off-Registry” IRR: 0.89, 95% CI:0.87-0.90); however, this relative difference was more pronounced for those “on-Registry” during the COVID-19 second wave compared to the same quarter a year earlier (multivariable-adjusted “on-Registry” IRR: 0.72, 95% CI: 0.68-0.77; “off-Registry” IRR: 0.83, 95% CI:0.82-0.85). ACSC hospitalization rates were estimated to be lower during the COVID-19 first wave compared to the same previous year quarter. However, for those “on-Registry”, this relative difference was not statistically significant (multivariable-adjusted “on-Registry” IRR: 0.78, 95% CI: 0.54-1.12; “off-Registry” IRR: 0.67, 95% CI:0.60-0.74).

**Table 2.** Crude rates, and crude and adjusted incidence rate ratios (IRR) comparing ED utilization and ACSC hospitalizations between COVID-19 first wave (Q2 2020) and Q2 2019; ED utilization between COVID-19 second wave (Q4 2020) and Q4 2019.

|  | “on-Registry” |  | “off-Registry” |  |
| --- | --- | --- | --- | --- |
|  | Rate/IRR | 95% C.I. | Rate/IRR | 95% C.I. |
| **COVID-19 first wave (Q2 2020) to Q2 2019** |  |  |  |  |
| **ED Utilization** |  |  |  |  |
| Rate for Q2 2020 | 117.9 |  | 86.2 |  |
| Rate for Q2 2019 | 150.1 |  | 101.0 |  |
| Crude IRR | 0.79 | 0.74-0.84 | 0.86 | 0.84-0.87 |
| Age-/Sex-adjusted IRR | 0.79 | 0.74-0.84 | 0.86 | 0.84-0.87 |
| Multivariable-adjusted* IRR | 0.86 | 0.81-0.92 | 0.89 | 0.87-0.90 |
| **ACSC Hospitalizations** |  |  |  |  |
| Rate for Q2 2020 | 13.8 |  | 8.7 |  |
| Rate for Q2 2019 | 20.6 |  | 14.8 |  |
| Crude IRR | 0.67 | 0.46-0.96 | 0.59 | 0.54-0.65 |
| Age-/Sex-adjusted IRR | 0.65 | 0.45-0.93 | 0.61 | 0.56-0.68 |
| Multivariable-adjusted* IRR | 0.78 | 0.54-1.12 | 0.67 | 0.60-0.74 |
| **COVID-19 second wave (Q4 2020) to Q4 2019** |  |  |  |  |
| **ED Utilization** |  |  |  |  |
| Rate for Q4 2020 | 103.6 |  | 83.0 |  |
| Rate for Q4 2019 | 154.4 |  | 105.5 |  |
| Crude IRR | 0.67 | 0.63-0.72 | 0.79 | 0.78-0.81 |
| Age-/Sex-adjusted IRR | 0.67 | 0.62-0.71 | 0.79 | 0.78-0.81 |
| Multivariable-adjusted* IRR | 0.72 | 0.68-0.77 | 0.83 | 0.82-0.85 |

\*Multivariable-adjusted: age, sex, Charlson index, rurality, census-level household income, CIMD composite index; (Rate per 1000 for ED contacts; Rate per 10,000 for ACSC hospitalizations)

### Interpretation

Although ED utilization decreased since the beginning of 2017, individuals “on-Registry” have substantially higher use of EDs than those “off-Registry”. ACSC hospitalizations were also higher for those “off-Registry” for multiple quarters. Rates of ED use and ACSC hospitalizations were lowest during NS’s first waves of COVID-19 for both those “on-” and “off-Registry.” A larger discrepancy in ED utilization between those “on-” and “off-Registry” was observed among individuals younger than 65 years. Females had higher rates of ED utilization, but ED use was similarly higher for “on-Registry” users, regardless of sex. While females did not exhibit notable differences by registry status, males who were “on-Registry” had somewhat higher rates of ACSC hospitalizations. Those “on-Registry” younger than 65 years had higher rates during the first wave of COVID-19. Compared to the analogous quarters a year earlier, ED utilization and ACSC hospitalizations were reduced during COVID-19, though for ACSC hospitalizations among those “off-Registry”, this difference was not statistically significant.

We cannot draw definitive conclusions about the rationale for decreased ED use during the pandemic. Patients may have been hesitant to use ED services out of fear of exposure, forgoing or receiving care elsewhere to reduce burden on EDs.(11,24) Instances of foregone care will have corresponding impacts on service use. Regarding sex-based differences, there is an abundance of research to suggest that women tend to use health services more than men,(25,26) which may contribute to higher ED use among females, if primary care services were being sought within the ED. Regardless, multiple Canadian studies have found that females are more likely to be frequent users of EDs.(27–29) Consistent with our findings, Canadian data indicate that males and older people have a greater number of ACSC hospitalizations than females and younger people, respectively.(30–32). This was true for older people “on-” and “off-Registry”, suggesting that current primary care models may be less effective in avoiding these types of admissions, regardless of attachment status. People ≥65 years are likely to be living with a chronic condition (33), thus it is plausible that older people require more urgent care, make fewer “discretionary” ED visits, and experience a higher number of ACSC. There are relatively few ACSC hospitalizations among the younger cohort, limiting inference. Though none have assessed health service utilization by attachment status or proxy (i.e., registration on centralized waitlist), our findings are consistent with other studies that have examined ED use and hospitalizations during the COVID-19 pandemic. These have shown marked decreases in ED use during waves of COVID-19.(34–36) In Alberta, ED visits for any reason decreased to 65% (IRR): 0.65, 95%CI: 0.62-0.67) and those for ACSC to 75% (IRR 0.75, 95%CI 0.72-0.79) compared to the previous year period.(36)

### Future directions

Our analyses quantify trends in ED use and ACSC hospitalizations; however, qualitative interviews may explain why these trends were found. As part of the PUPPY study(14), we have interviewed healthcare providers and knowledge users. Initial evidence provides mixed support for our findings; some themes support decreased ED use, including patient fear and provider reluctance to send patients to the ED to avoid overcapacity; others describe experiences that may have motivated increased ED utilization, such as patients misunderstanding COVID restrictions and providers sending patients to the ED when they could not see them themselves. Patient interviews planned as part of the PUPPY study will contribute to understanding where patients accessed care during the pandemic and any health implications associated with these decisions. During the pandemic, there were policy changes and innovations to help maintain primary care access for patients, including increases in the provision of virtual care.(37) Patients experienced delays accessing primary care,(38,39) influencing the need to visit the ED, regardless of attachment status. Future studies could explore the frequency of virtual care access by patient attachment status.

### Limitations

We cannot definitively determine to what extent observed trends are due to the COVID-19 pandemic or attachment. Further, while the comparison of COVID-19 waves with analogous calendar periods the prior year may effectively adjust for seasonality in the estimation of rate ratios, there may be important unmeasured confounders that were unaccounted for in the analyses. For one, the Canadian Index of Multiple Deprivation may not exclusively capture important variation in socioeconomic need, which might have undermined our ability to control for the impact of related factors on differences in health service utilization outcomes during COVID-19 compared with the prior-year period. More precise socioeconomic measures will further inform the impact of pandemics on access to health services and how this differs across those actively seeking attachment. We did not adjust for time on waitlist, which may indicate increased need or deprivation of care. We could not verify that all centralized waitlist users were identified in the HDNS Insured Patient Registry, though we have no reason to believe that our “on-Registry” sample, which captured the majority, does not represent the Registry user base. We were, however, unable to account for those unattached who were not on the Registry, introducing misclassification in the interpretation of registry status as an attachment proxy. This study may have been limited by using a physician billing database to enumerate ED use. This database only captures ED visits where a physician assessed the patient and submitted a billing claim.

## Conclusion

Access to primary care is essential for preventative care, population health outcomes, and reducing the acute care burden. Nova Scotians actively seeking primary care attachment utilize non-critical hospital-based services more frequently. However, those on and off the waiting list for primary care provider attachment had lower utilization of these services during the COVID-19 pandemic. The degree to which forgone services produce downstream health burden remains to be seen, the preliminary assessment of which is part of ongoing PUPPY study research.

## Data Availability

The data presented in this study are not publicly available. The data were obtained from Health Data Nova Scotia through a data sharing agreement with the Nova Scotia Department of Health and Wellness and Nova Scotia Health. They were accessed by the authors through a formalized process established by the aforementioned parties. Others may request access to these data by contacting the Health Data Nova Scotia directly and submitting a data access request.

## Declarations

Authors have no conflicts of interest to declare.

## Data sharing

Some of the data presented in this study are not publicly available. The data were obtained from Health Data Nova Scotia through a data sharing agreement with the Nova Scotia Department of Health and Wellness and Nova Scotia Health. They were accessed by the authors through a formalized process established by the aforementioned parties. Others may request access to these data by contacting the Health Data Nova Scotia directly and submitting a data access request.

## Acknowledgements

The data used in this report were made available by Health Data Nova Scotia of Dalhousie University. Although this research is based on data obtained from the Nova Scotia Department of Health and Wellness, the observations and opinions expressed are those of the authors and do not represent those of either Health Data Nova Scotia or the Department of Health and Wellness.

